# Rethinking respiratory disease forecasting: temporal heterogeneity between surveillance predictors and outcomes drives forecast instability

**DOI:** 10.64898/2026.08.19.26360833

**Authors:** Hillary M. Topazian, Theresa R. Sheets, Randon J. Gruninger, Joshua Kelley, Nathan LaCross, Matthew H. Samore, Eric Lofgren, Lindsay T. Keegan

## Abstract

Since the COVID-19 pandemic, forecasting hubs and non-traditional respiratory disease surveillance streams have become increasingly common. However, many forecasting approaches assume that relationships between surveillance predictors and disease outcomes remain stable over time and that incorporating additional historical data will improve forecast performance. To evaluate these assumptions in a real-world setting, we developed and evaluated forecasts of SARS-CoV-2 and influenza hospitalizations in Utah using syndromic surveillance, test positivity, and wastewater data. Rather than identifying a single, best-performing model, we examined whether relationships between surveillance predictors and hospitalization outcomes remained stable across seasons and whether longer historical training periods consistently improved forecast accuracy. Relationships between surveillance predictors and hospitalizations varied substantially by pathogen and season. Analyses using pooled data across multiple years suggested strong positive correlations between predictors and outcomes, but these aggregated patterns often obscured weak or negative correlations observed during SARS-CoV-2 variant waves and influenza seasons. Forecast performance similarly varied over time. Models that performed well during some seasons, transmission phases, or under certain training strategies frequently performed worse than benchmark models in others. Training on additional historical data generally reduced forecast accuracy, though this varied by disease and transmission phase. Forecasting groups should prioritize continual evaluation of surveillance predictors, adaptive strategies, and diverse ensembles, rather than relying on a single model, data stream, or historical training framework each year.

**AUTHOR SUMMARY:** Respiratory disease forecasting hubs and novel data streams have become integral parts of infectious disease surveillance and public health decision-making since the COVID-19 pandemic. Many forecasting groups assume that adding more historical data will improve model performance and that relationships between surveillance predictors, such as emergency department visits or wastewater, and hospitalizations will remain stable over time. We evaluated these assumptions using forecasts of SARS-CoV-2 and influenza hospitalizations in Utah. We found that relationships between surveillance predictors and hospitalizations varied across SARS-CoV-2 variants, influenza seasons, and periods of increasing and decreasing transmission. Forecast performance also varied considerably, with models that performed well in some seasons often performing poorly in others. Public health groups should continually evaluate the utility of surveillance predictors in real-time and prioritize adaptable, diverse modeling approaches.

## INTRODUCTION

Seasonal respiratory viruses, including SARS-CoV-2 and influenza, impose a large and highly variable burden of disease, hospitalizations, and deaths each year. In the 2024-2025 season alone, there were an estimated 1.2 million to 2.2 million respiratory disease hospitalizations in the United States [1–3]. Severe seasons of a single virus or overlapping peaks across multiple viruses can overwhelm the capacity of emergency departments (ED) and hospitals. This is of particular concern with the occurrence of “triple-demic” outbreaks of SARS-CoV-2, influenza, and respiratory syncytial virus (RSV) [4], and as vaccination rates have fallen in the 2025-2026 season in response to mixed messaging and changing policies [5,6].

The infectious disease surveillance landscape has undergone a rapid transformation in recent years. The COVID-19 pandemic accelerated the growth of non-traditional data steams such as ED syndromic surveillance and wastewater surveillance [7], yet the relative value of these inputs remains unknown. At the national level, modeling teams submitting forecasts to the Centers for Disease Control and Prevention’s (CDC) modeling hubs generally train on weekly cases, hospitalizations, and deaths, and are often unable to incorporate high-resolution covariates, such as age-stratified syndromic surveillance, and wastewater viral concentrations, because these data are not consistently available across all 50 states [8,9]. It remains unclear which data streams improve forecast accuracy, how model types behave under these shifting data streams, and whether the predictive power of covariates differs across time and pathogens.

As forecasting systems increasingly seek to incorporate novel data streams, understanding how their predictive value changes over time becomes important. Accurate forecasting at the national, state, and local levels can inform health system decision-making to manage surges, such as promoting vaccination, enacting social distancing measures, and adjusting staffing schedules at local hospitals [10,11]. While forecasting hubs are increasingly used to support public health planning and hospital preparedness [12], many approaches rely on the assumption known as “stationarity”, that historical relationships between surveillance predictors and disease outcomes remain relatively stable over time [13]. However, respiratory disease systems are fundamentally nonstationary: transmission dynamics, dominant strains and variants, population immunity, healthcare-seeking behavior, and surveillance practices all change over time [14,15]. These shifts may substantially alter the predictive utility of historical data and covariates across seasons and outbreaks. The fluctuating availability of data streams also leads to “surveillance drift”, such as during the COVID-19 pandemic where modelers initially relied on clinical testing early on in the pandemic, eventually transitioning to a focus on active and then passive hospital reporting, and finally incorporation of wastewater surveillance [7].

One consequence of the stationarity assumption is that many forecasts continue to rely on long training windows under the assumption that incorporating additional years of data could improve predictive performance [16–18]. Existing respiratory disease modeling hubs, such as the CDC’s CovidHub [12], FluSight [19], RSVHub [20], and Flu MetroCast Hub [21] initiatives, often use models trained on pooled historical data and weighted ensemble approaches which likewise assume that historically well-performing models will remain reliable contributors as disease activity shifts across phases of an epidemic curve through increasing, peak, and decreasing transmission [22]. However, if relationships between predictors and outcomes change over time, incorporating older data may instead degrade forecast accuracy by training on patterns that are no longer epidemiologically relevant.

To evaluate these assumptions in a real-world public health setting, we developed and evaluated forecasts of SARS-CoV-2 and influenza hospitalizations in Utah using multiple surveillance data streams over time. Rather than identifying a single optimal model, we examined whether relationships between surveillance data predictors and hospitalization outcomes remain stable between respiratory virus seasons, and whether incorporating longer historical training periods consistently improves predictive performance. We evaluated multiple model classes, surveillance streams, and training strategies across disease types and time periods. By characterizing how respiratory disease forecasts perform under changing conditions, we hope to better inform Utah’s seasonal prediction models to support timely state-level public health decision-making.

## METHODS

This study protocol was classified as non-research by Institutional Review Boards at the University of Utah and the Utah Department of Health and Human Services (UDHHS).

### Setting and data sources

To examine the temporal relationships between surveillance predictors and respiratory disease hospitalization counts, we focused our forecasting efforts on Utah. Utah has a population of approximately 3.5 million people (2024) and comprises 29 counties [23]. The population distribution of Utah is similar to many western and mid-western states, where the majority of the state’s population is centralized in a major metropolitan area and the remainder of the state is classified as rural [23]. Our long-standing close collaboration with UDHHS enabled sharing of data streams that would not otherwise be available. These data included age-stratified hospitalizations, age-stratified syndromic surveillance, SARS-CoV-2 testing, and wastewater viral concentrations, and are described below.

#### Hospitalizations

Daily new hospital admission counts were obtained for each respiratory infection stratified by age group: pediatric (0-17 years), adult (18-64 years), and older adult (65+ years). Hospitalization data have historically been collected through two mechanisms: 1) active surveillance, in which UDHHS investigators reach out to and directly collect data from hospitals, and 2) passive reporting, in which healthcare systems routinely submit data to UDHHS. During the height of the COVID-19 pandemic, SARS-CoV-2 data were collected actively; currently, hospitalization data for all respiratory infections, including influenza, are collected passively.

A SARS-CoV-2 or influenza hospitalization was defined as an inpatient admission with a positive result on a laboratory test collected from 14 days pre-admission through three days post-admission [24]. For this analysis, SARS-CoV-2 hospitalization data were available starting March 20, 2020 and influenza data from January 1, 2018. Data streams for all infections were available through May 1, 2026. Utah hospitals serve as tertiary centers for transfers of patients from neighboring states, including Idaho, Nevada, Montana, and Wyoming. Data for non-Utah residents (an estimated 5%-15%) were excluded from all data streams to capture the dynamics of within-state transmission.

#### Syndromic surveillance

The objective of this analysis was to identify predictors that precede changes in respiratory disease burden, and we incorporated ED syndromic surveillance data as an established early-warning measure of community respiratory virus activity. Syndromic surveillance is designed to support early detection of outbreaks by using routinely collected health-related information, including symptoms, clinical signs, and preliminary diagnoses, rather than relying exclusively on microbiologically or clinically confirmed cases [25,26]. These data include the number of ED patient encounters with a preliminary diagnosis of SARS-CoV-2 or influenza captured through either ICD-10 codes or classification as “COVID-like illness” and/or “influenza-like illness”. This analysis classifies encounters through ICD-10 codes to remain consistent with the National Syndromic Surveillance Program [27]. These data are reported daily by all EDs in the state and include both counts and percentages of total ED visits attributable to each respiratory infection. UDHHS receives data from all 49 EDs in Utah, resulting in 100% coverage of the population. Data are stratified by age group: pediatric (0-17 years), adult (18-64 years), and older adult (65+ years). For this analysis, SARS-CoV-2 syndromic surveillance data were available starting from March 20, 2020, and influenza data from January 1, 2018.

#### Test positivity

Throughout the COVID-19 pandemic, the test positivity rate was used as a proxy for the current level of transmission in a community [28] and often included in news reports as a measure of pandemic severity. We evaluated the predictive power of the SARS-CoV-2 test positivity data stream as an early indicator of COVID-19 hospitalizations in our setting as has been observed elsewhere [29]. Test positivity provides a measure of the proportion of reported tests resulting in confirmed infection and may reflect changes in transmission intensity, healthcare-seeking behavior, and testing availability. Testing data were available for SARS-CoV-2 only, from March 6, 2020 to January 14, 2025, after which testing centers began closing. Test positivity was defined as the number of positive tests over the total number of tests per week. Both RT-PCR amplification tests and rapid antigen test results were included if conducted in a healthcare setting; at-home rapid antigen tests were not reportable.

#### Wastewater surveillance

To capture community-level respiratory virus activity independent of clinical testing patterns, we incorporated wastewater surveillance data from Utah’s statewide wastewater monitoring program. Wastewater data provide sewershed-level measurements of the presence and concentration of SARS-CoV-2 and influenza in the wastewater. Viral concentrations were estimated from samples collected at participating wastewater treatment facilities, quantified via droplet digital PCR, and normalized by wastewater flow during the sampling period and by the estimated sewershed population [30]. The output of interest was the daily concentration in million gene copies per person per sewershed per day (MCG/person/day). To facilitate comparison across the study period, all MGC values were divided into deciles based on the full distribution, and each day was assigned a corresponding decile rank.

Utah began reliably collecting wastewater data for SARS-CoV-2 on May 1, 2020 and for influenza on November 13, 2024. Influenza wastewater concentration values were grouped by subtype A and subtype B (**Figure S1**). Type A values alone were used in this analysis as A and B assays are not directly comparable and influenza A predominates in the United States each season [31].

During the study period, 35 wastewater treatment facilities across the state contributed data, covering approximately 88% of Utah’s population [30]. Routine sampling occurred roughly twice a week on Tuesdays and Thursdays, and most participating utilities collected either flow- or time-weighted composite samples over approximately 24-hours. Two facilities collected 6-hour manual composite samples, and one facility collected grab samples. Since hospitalization and ED data were reported daily, we assigned daily wastewater values by carrying forward the most recent value on days missing a sample collection (e.g. Monday, Wednesdays). Weights to aggregate sewershed level data to the state level were created by assigning all address points in Utah to a sewershed and calculating the proportion of addresses within each block. Address points were pulled from the Utah Geospatial Resource Center [32]. Sewershed boundaries were contributed by UDHHS.

Concentrations below the reporting limit were set to half of the reporting limit value (SARS-CoV-2: 7.89 gene copies/mL wastewater; influenza A: 9.68 gene copies/mL wastewater) prior to normalization by flow and population. As data were collected at the sewershed level, values are highly variable and susceptible to seasonal temperature fluctuations, chemical contamination, and rainfall, all of which affect RNA amplification and effluent concentration. It was also impossible to separate the contributions of local residents from those of commuters and tourists. Nevertheless, wastewater has been successfully used to forecast respiratory disease in a number of localities [33].

### Covariate predictor selection and lag assessment

To identify surveillance predictors that could provide early information about subsequent respiratory disease hospitalizations, we first evaluated both the short-term and long-term temporal relationships between potential predictors for COVID-19 and influenza.

This screening step was used to characterize which data streams were most strongly associated with hospitalizations, whether those associations varied over time, and whether candidate predictors tended to lead or lag hospitalizations. Candidate predictors included age-stratified hospitalizations, age-stratified ED counts and percentages of total encounters (syndromic surveillance metrics), wastewater viral concentrations categories, and test positivity. Test positivity was evaluated for SARS-CoV-2 only because comparable testing data were not available for influenza. Covariate data were aggregated from the daily to the weekly level to harmonize the temporal resolution of data streams and to reduce noise in daily reporting. Daily count variables, including hospitalizations and ED encounters were summed by week. Wastewater concentrations and SARS-CoV-2 test positivity values were summarized using a weekly median.

To determine the correlation between variables, we computed pairwise associations of variables using Spearman’s rank correlation coefficient on complete-case observations. To reduce spurious correlations driven by shared epidemic trajectories, seasonality, or long-term changes in reporting, each variable was detrended separately within each pathogen and variant period (or influenza season). Detrending was performed by fitting a generalized additive model (GAM) with a smooth function of time and using the resulting residuals for subsequent correlation analyses.

To quantify short-term temporal correlation between variables, we evaluated lead-lag relationships between candidate predictors and all-age hospitalizations using cross-correlation analysis. For each pathogen, cross-correlations were calculated across lags ranging from -6 to +6 weeks. Positive lags indicated that the predictor led hospitalizations while negative lags indicated the predictor lagged hospitalizations.

Cross-correlations were calculated using detrended residuals to emphasize short-term lead-lag relationships independent of the overall epidemic trajectory and were restricted to complete observations.

To characterize the seasonal (influenza) or variant-dependent (SARS-CoV-2) predictive power of the covariates, we stratified influenza data by season, with each season beginning July 1 and running for one year; and disaggregated COVID-19 data by dominant variant type using sequencing data provided by UDHHS (**Figure S2**).

### Models

We evaluated 20 models to forecast daily hospitalization counts for each respiratory disease (**Table S1**). These models included two benchmark models, two statistical time series models, one semi-mechanistic decomposable regression model, nine flexible regression models, and six mechanistic simulation-based models.

Benchmark models included a naïve model, which projects the most recently observed value forward, and a drift model, which extends the naïve model by incorporating a linear trend based on the average change from the first to the most recent observation. Statistical time series models included Auto Regressive Integrated Moving Average (ARIMA) models and exponential smoothing (or Error, Trend and Seasonality, ETS) models. ARIMA models use lagged observations, differencing, and error terms to capture temporal dependence, while ETS models decompose series into error, trend, and seasonal components updated via exponential smoothing. The semi-mechanistic model was Prophet, developed by Meta, which is a decomposable time series approach with piecewise linear or logistic trends which can accommodate multiple seasonalities, change points, and holiday effects [34]. Flexible regression models included GAMs, which use spline-based non-linear smoothing functions to model effects of time and covariates. GAM models incorporated syndromic surveillance and wastewater covariates. Mechanistic simulation-based models included Mantis, a foundation model developed at the University of Michigan and trained on mechanistic simulations generalized across diseases and disease outcomes [35]. Covariates from syndromic surveillance and wastewater data were incorporated into Mantis.

Additional model details are provided in the Supporting Information.

### Training and forecasting

Individual models were trained on observed daily hospitalization counts for either a fixed window of 12 weeks or for a telescoping (increasing) window which expanded to include all available historical data. All models used a 3-week test period. **Figure 1** illustrates our forecasting framework. To incorporate syndromic surveillance and wastewater covariates into hospitalization forecasts, these predictors were first projected forward at a daily level using GAM models with spline-based smooth functions of time. Forecasted covariate values were then used as inputs to downstream hospitalization forecasting models.

**Figure 1.**
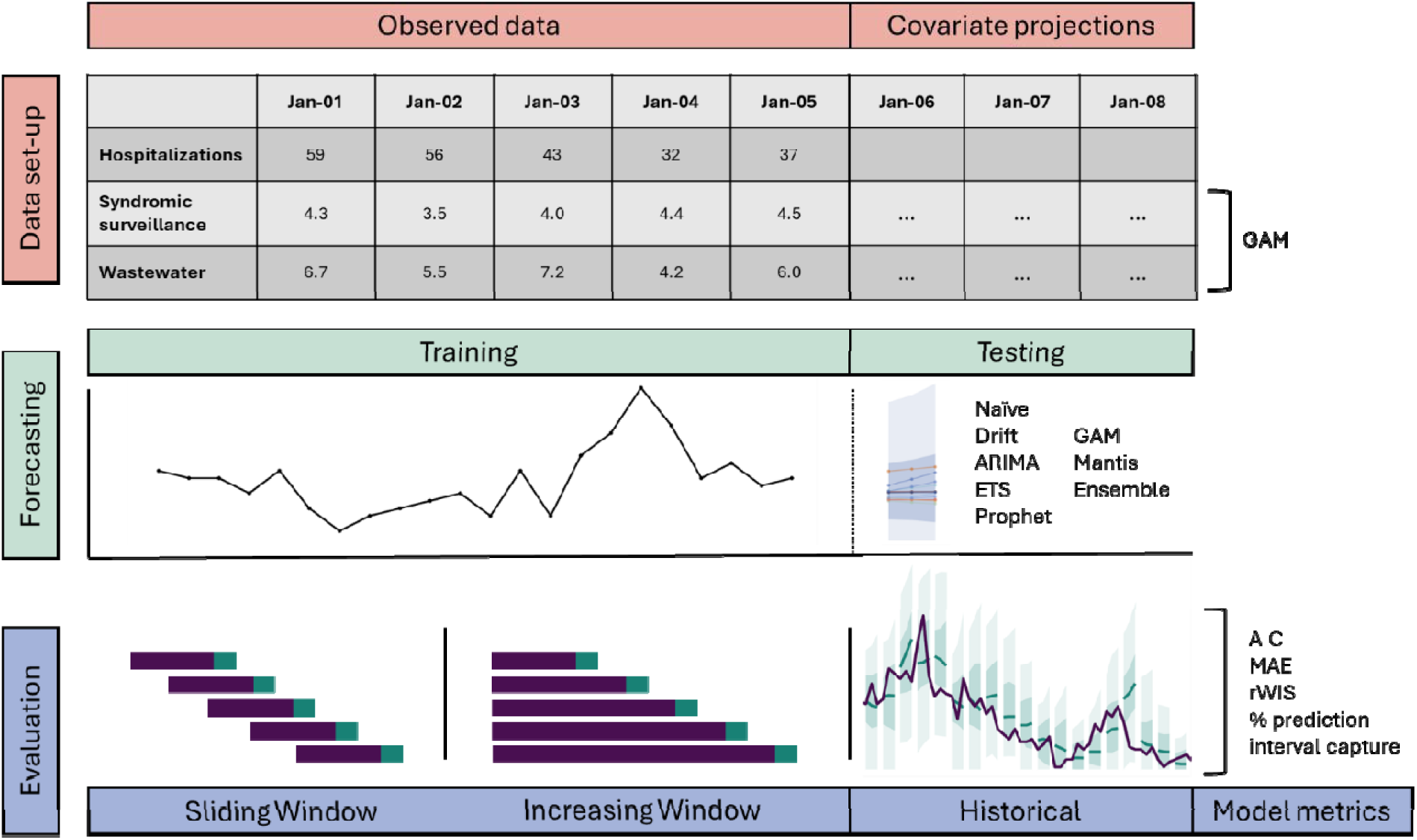
Diagram of forecasting methodology. <u>Data set-up</u>: observed data for hospitalizations and other predictors were classified as one record per day. Covariate values for wastewater and syndromic surveillance were projected into the testing period using generalized additive models (GAMs). <u>Forecasting</u>: models were trained on observed daily hospitalization counts and forecasts were output for a three-week testing period. Twenty individual models were used to generate an ensemble model forecast. <u>Evaluation</u>: a sliding window and increasing window training approach were run for each respiratory infection and compared. Individual model fit and model performance were also assessed over time using the Akaike Information Criterion (AIC), Mean Absolute Error (MAE), relative weighted interval score (rWIS), and the percent of forecasts captured within the 50^th^ and 95^th^ percentile prediction intervals. Note: ARIMA = Autoregressive Integrated Moving Average; ETS = Error, Trend, Seasonality

Due to significant weekly periodicity in hospitalization data, with a greater number of admissions occurring on Mondays compared to other days of the week (SARS-CoV-2: 15.8%, influenza: 15.1%), and lower counts on Saturdays and Sundays, model runs were initialized on Thursdays to maintain consistent weekly alignments (**Figure S3**). All models were fit using daily data, except for Mantis models which operate only at the weekly level. For ease of comparison, we aggregated all model results to the weekly level when assessing model fit, accuracy, and visualizing results. Daily forecasts were used by UDHHS for internal planning.

Ensemble forecasts were constructed by taking the median predicted hospitalization counts across the 20 individual models at each time step and for each disease type and model type. Corresponding 50% and 95% prediction intervals were also defined as the median value of model-specific intervals at each time step.

All analyses were conducted in R 4.5.2 (R Core Team, 2026). Forecasting models were run using the *forecast* (*v8.24.0*; Hyndman and Khandakar, 2025), *mgcv* (*v1.9-3*; Wood, 2025), *prophet* (*v1.0*; Taylor and Letham, 2021), and *rmantis* (*v0.1.0*; Dudley and Magdaleno, 2026) packages. Code for data manipulation, analysis, and visualizations can be found at https://github.com/EpiForeSITE/respiratory-forecasting-Utah.

### Model evaluation

Model fit was assessed using the Akaike Information Criterion [36] (AIC) for likelihood-based models. Forecasting accuracy was assessed using the relative weighted interval score (rWIS), which scales the weighted interval score (WIS) of each model by that of the naïve benchmark model [37]. WIS evaluates forecasts by combining penalties for prediction interval width, overprediction, and underprediction. The percentage of forecasts captured in the 50% and 95% prediction intervals were also reported by pathogen and by time period. In addition to assessing model performance over an entire season, accurately predicting an inflection point (peak or trough) is a key challenge for respiratory virus forecasting. A sub-analysis of rWIS during 10 COVID-19 peaks and 9 troughs and 7 influenza peaks and 7 troughs was also performed to assess model performance during inflection points. Each peak and trough represented a three-week period (**Figure S4**).

## RESULTS

### Outbreak Characteristics

Longitudinal respiratory disease data are shown in **Figure 2**. COVID-19 hospitalization and ED values were highest during the pandemic period (June 2020 – February 2022), reaching a peak of 651 all-age hospitalizations per week and 18.8% of all-age ED visits, before converging to steady state dynamics as a year-round virus with waves driven by the emergence of new variants. Influenza followed cyclical seasonal trends; however, the onset of high transmission waves and peak intensity varied from year to year. The highest recorded weekly influenza hospitalization count was 333 in 2025. Wastewater decile data tracked alongside hospitalization and ED visit trends. High viral concentrations were still detected for SARS-CoV-2 in years following the pandemic, despite the lower number of hospitalizations and ED visits recorded. Age-stratified hospitalization and ED trends are shown in **Figure S5**.

**Figure 2.**
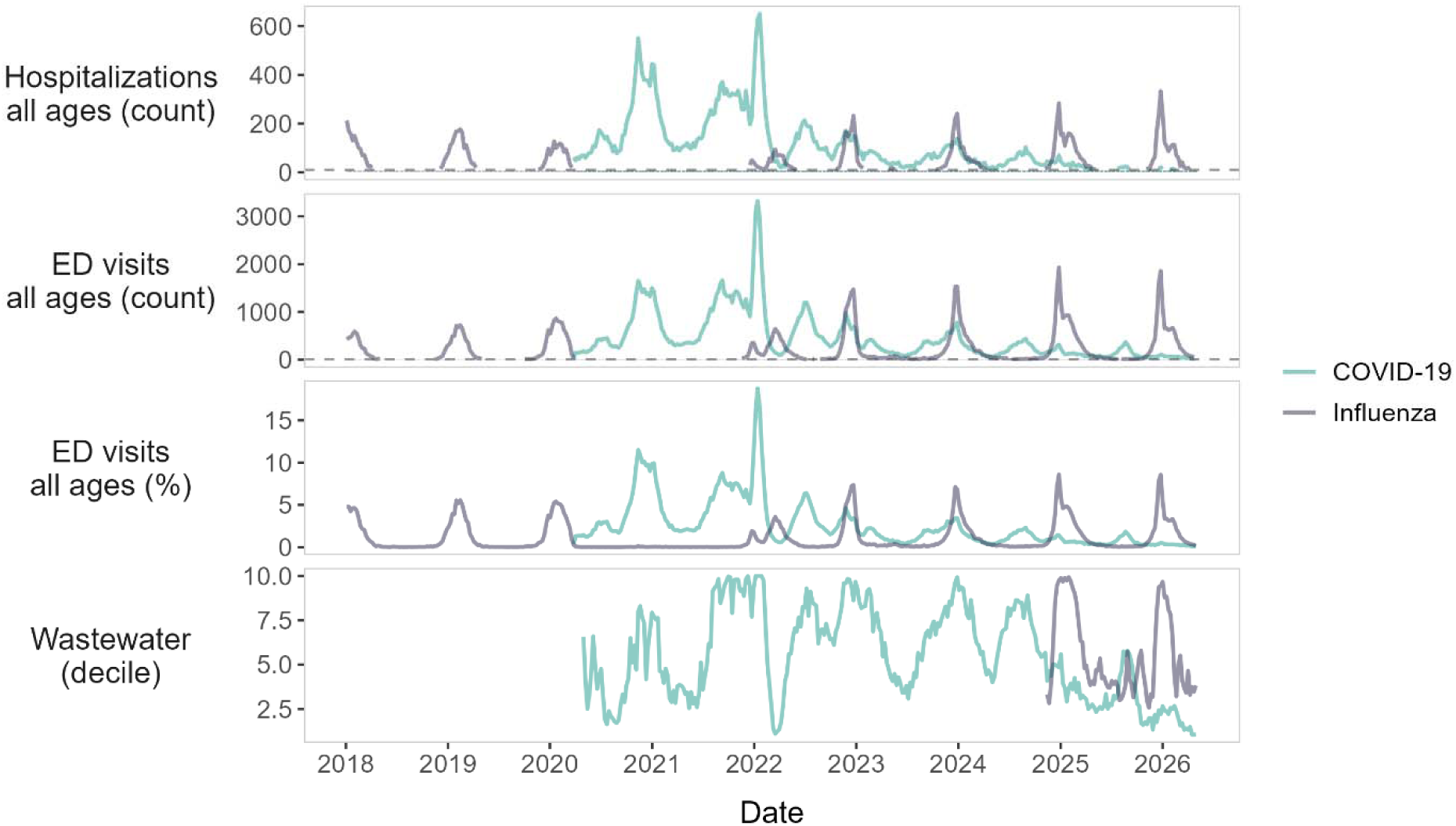
Data streams. Weekly time series of each data stream and selected predictors used in forecasting models. Counts of hospitalizations and syndromic emergency department visits (ED) between 1 and 10 per week are censored to protect patient confidentiality. Age-stratified hospitalization and ED trends are shown in **Figure S5**.

### Covariate predictive relationships

A cross-correlation analysis of age-stratified hospitalization, ED, wastewater, and test positivity variables demonstrated substantial variability in predictor relationships across disease type, age group, and time. Across all years of COVID-19 data, adult hospitalizations, adults ED visits, and wastewater measures exhibited a strong positive correlation (**Figure 3**). Conversely, pediatric hospitalizations, pediatric ED visits, and test positivity measures were generally weakly correlated with other variables. When all years of data were combined, all influenza predictors, apart from wastewater, showed consistent strong positive correlations across variables and age groups.

**Figure 3.**
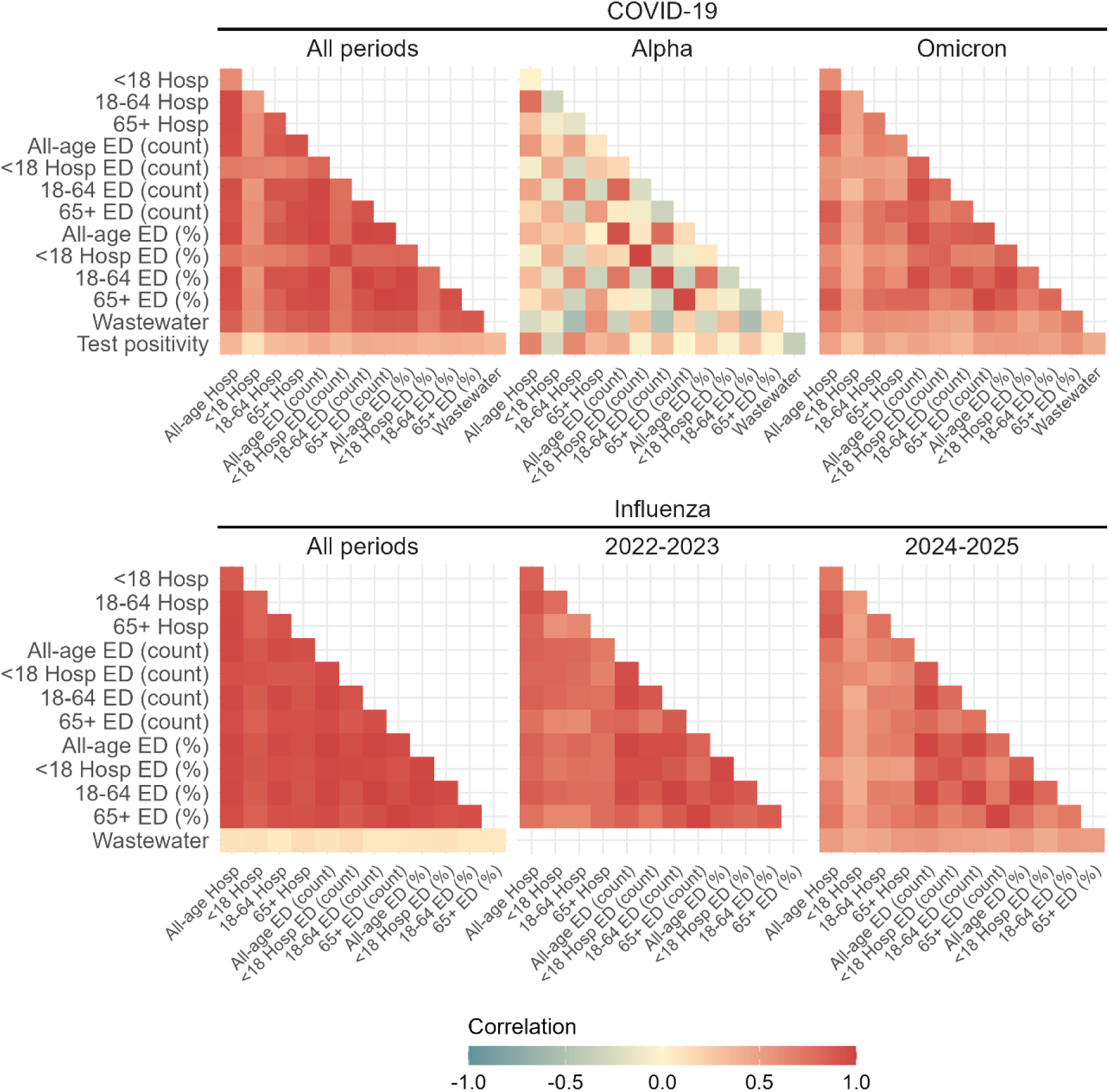
Predictor cross-correlations. Cross-correlation matrices of age-stratified hospitalization, emergency department (ED), and wastewater surveillance predictors for COVID-19 and influenza across all periods and by season or dominant variant. Warmer colors indicate stronger positive correlations, while cooler colors indicate weaker or negative correlations. <18 refers to pediatric hospitalizations under 18 years of age, 18-64 refers to adult hospitalizations, and +65 refers to hospitalizations among adults 65 years of age and older. Results stratified by all COVID-19 variant periods and influenza seasons are shown in **Figure S6**.

However, when cross-correlation analyses were stratified by longer-term temporal trends, dominant-variant period for COVID-19, and season for influenza, results differed considerably. For COVID-19, few predictors were correlated, with pediatric hospitalizations, ED, and wastewater predictors negatively correlated with predictors from older age groups during the Alpha wave in 2021. Conversely, the Omicron wave and the current COVID-19 variant demonstrated stronger positive correlations among adult hospitalization, ED, and wastewater variables (selection of time periods shown in **Figure 3**, all time periods shown in **Figure S6**). Influenza also demonstrated between-season variability in predictor correlations, but patterns were generally more consistent than those observed for COVID-19.

Correlations between age-stratified variables and all-age hospitalizations counts showed broadly similar patterns when variables were shifted from 6 weeks leading to 6 weeks lagging all-age hospitalization outcomes (selected time periods shown in **Figure 4**, all time periods shown in **Figure S7)**. Across all years of COVID-19 data, adult hospitalization, ED, and wastewater variables remained strongly correlated with all-age hospitalization counts, even when lagged by several weeks. When stratified by dominant variant period, similar relationships were observed during periods dominated by the original COVID-19 strain, Delta, Omicron, and the current variant, although the strongest correlations generally occurred with no lag time. Few correlations between variables and all-age hospitalizations were observed during time periods when Alpha dominated. Influenza variables showed similar patterns, but strongly positive correlations existed across all age strata and were limited to within 2 weeks of lead or lag time.

**Figure 4.**
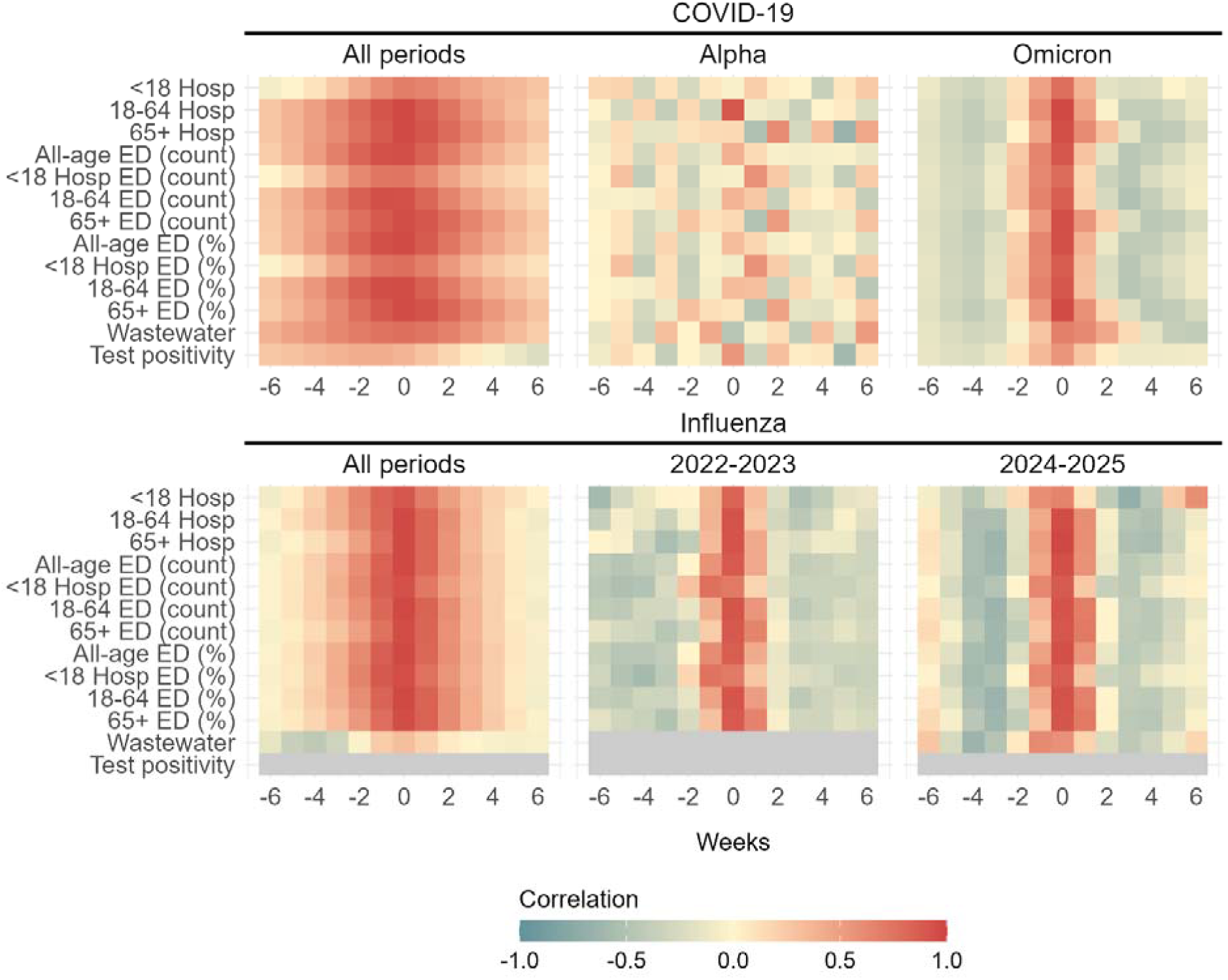
Lagged cross-correlations. Lagged correlations between all-age hospitalization counts and hospitalization, emergency department (ED) surveillance, wastewater, and test positivity (SARS-CoV-2 only) predictors. Results are stratified by COVID-19 and influenza across all periods and by season or dominant variant. Correlations are shown across weekly lags (-6 to +6 weeks), where negative lags indicate that predictors lag hospitalizations and positive lags indicate that predictors lead hospitalizations. Warmer colors indicate stronger positive correlations, while cooler colors indicate weaker or negative correlations. <18 refers to pediatric hospitalizations under 18 years of age, 18-64 refers to adult hospitalizations, and +65 refers to hospitalizations among adults 65 years of age and older. Results stratified by all COVID-19 variant periods and influenza seasons are shown in **Figure S7**.

### Model fit

Median AIC values were generally lower for GAM models than for benchmark models, indicating that added flexibility and covariate inclusion improved model fit (**Figure S8**). ETS models had the highest median AIC values across all COVID-19 variants and influenza seasons, and GAM models had the lowest AIC values across all time periods for all likelihood-based models.

Forecasts run under the sliding window training strategy generally resulted in higher 50% and 95% prediction interval coverage than forecasts run under the increasing window training strategy for both COVID-19 and influenza (**Figure 5**, **Figure S9**).

**Figure 5.**
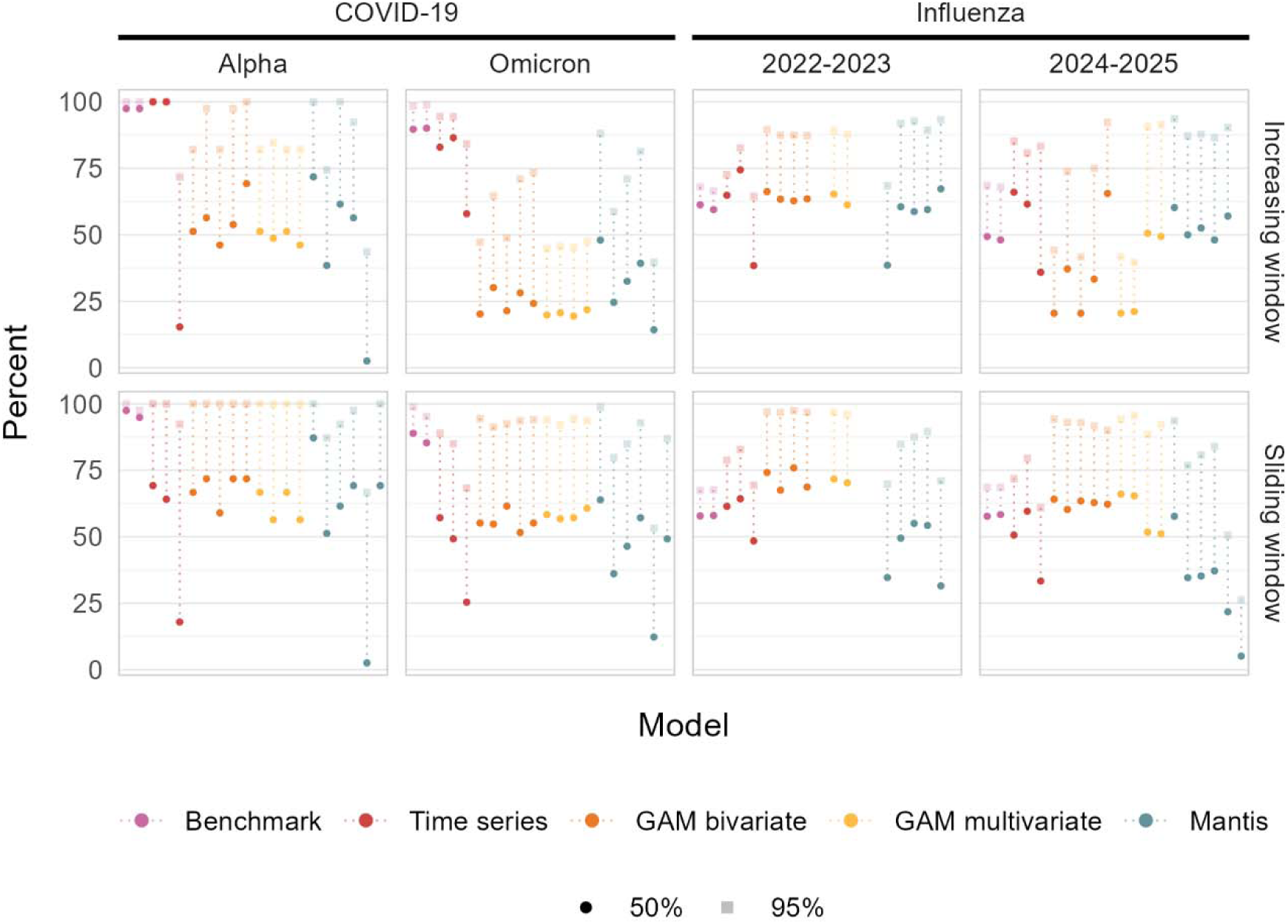
Prediction interval coverage. Prediction interval metrics (50% and 95%) for respiratory disease forecasts, stratified by season and time-series training method. Results from all COVID-19 variant periods and influenza seasons are shown in **Figure S9**. Note: GAM = generalized additive model rWIS metrics also revealed that the sliding window training strategy outperformed the benchmark models for most COVID-19 and influenza models, with median rWIS values under 1 (Figure 6, **Figure S10**). Under the increasing window strategy, GAM model performance fluctuated across time periods. rWIS also varied by dominant-variant: most GAM models achieved rWIS below 1 during the Alpha, Delta, and current COVID-19 variant periods, as well as during the 2022-2023 influenza season; whereas median rWIS values were far greater than 1 under the increasing window strategy for many GAM models during the Omicron period and the 2021-2022, 2023-2024, and 2023-2025 influenza seasons, indicating that there may be temporal trends that hinder model performance. Mantis models were robust to training strategy, generally outperforming benchmark models across most time periods. Evaluation of median absolute errors showed similar trends (**Figure S11**).

Increasing window forecasts only outperformed sliding window forecasts during the 2022-2023 influenza season where 77% of forecasts had higher 50% prediction interval coverage.

### Model fit at inflection points

An analysis of 10 COVID-19 peaks and 9 troughs and 7 influenza peaks and 7 troughs demonstrated poor model reliability during major transmission change points. During COVID-19 hospitalization peaks, 39% of forecasts under the increasing window strategy had rWIS values greater than 1, compared with 33% of forecasts under the sliding window strategy (**Figure 7**). Influenza forecasts performed even more poorly during peaks, with rWIS values greater than 1 in 67% of forecasts under the increasing window strategy and 73% under the sliding window strategy. Model performance was improved during trough periods when transmission begins to exponentially increase; across measured troughs, the proportion of forecasts with rWIS values greater than 1 was 33% under the increasing window strategy and 9% under the sliding window strategy for COVID-19 forecasts. For influenza troughs, this proportion was 23% under the increasing window strategy and 40% under the sliding window strategy.

**Figure 6.**
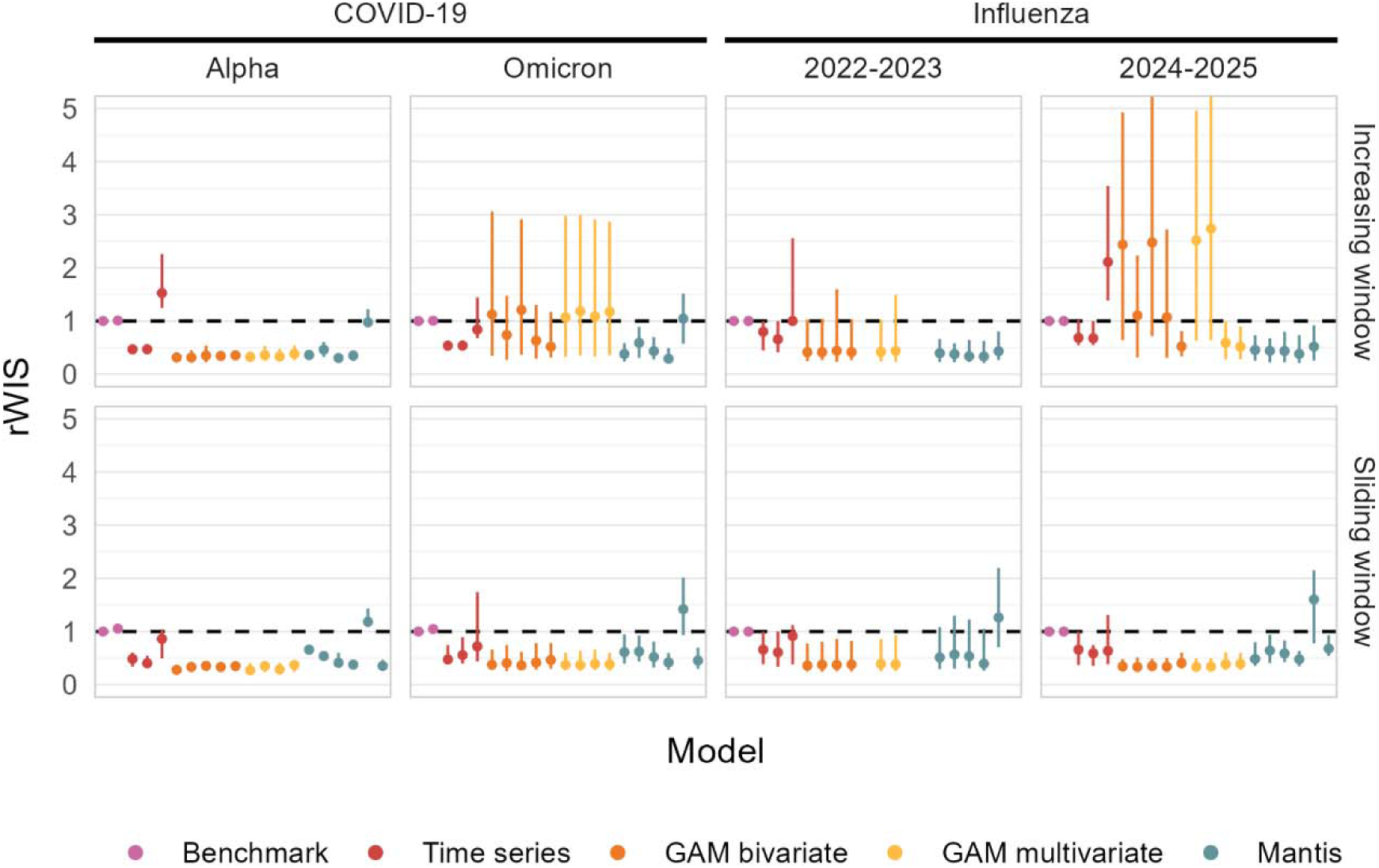
Relative weighted interval score. Median (interquartile range) relative weighted interval score (rWIS) metrics, stratified by season and time-series training strategy. The dashed horizontal line indicates an rWIS value of 1, relative to a Naïve benchmark (a lower score = better performance). Infinite values due to benchmark model WIS = 0 have been removed. Results from all COVID-19 variant periods and influenza seasons are shown in **Figure S10**. Note: GAM = generalized additive model

**Figure 7.**
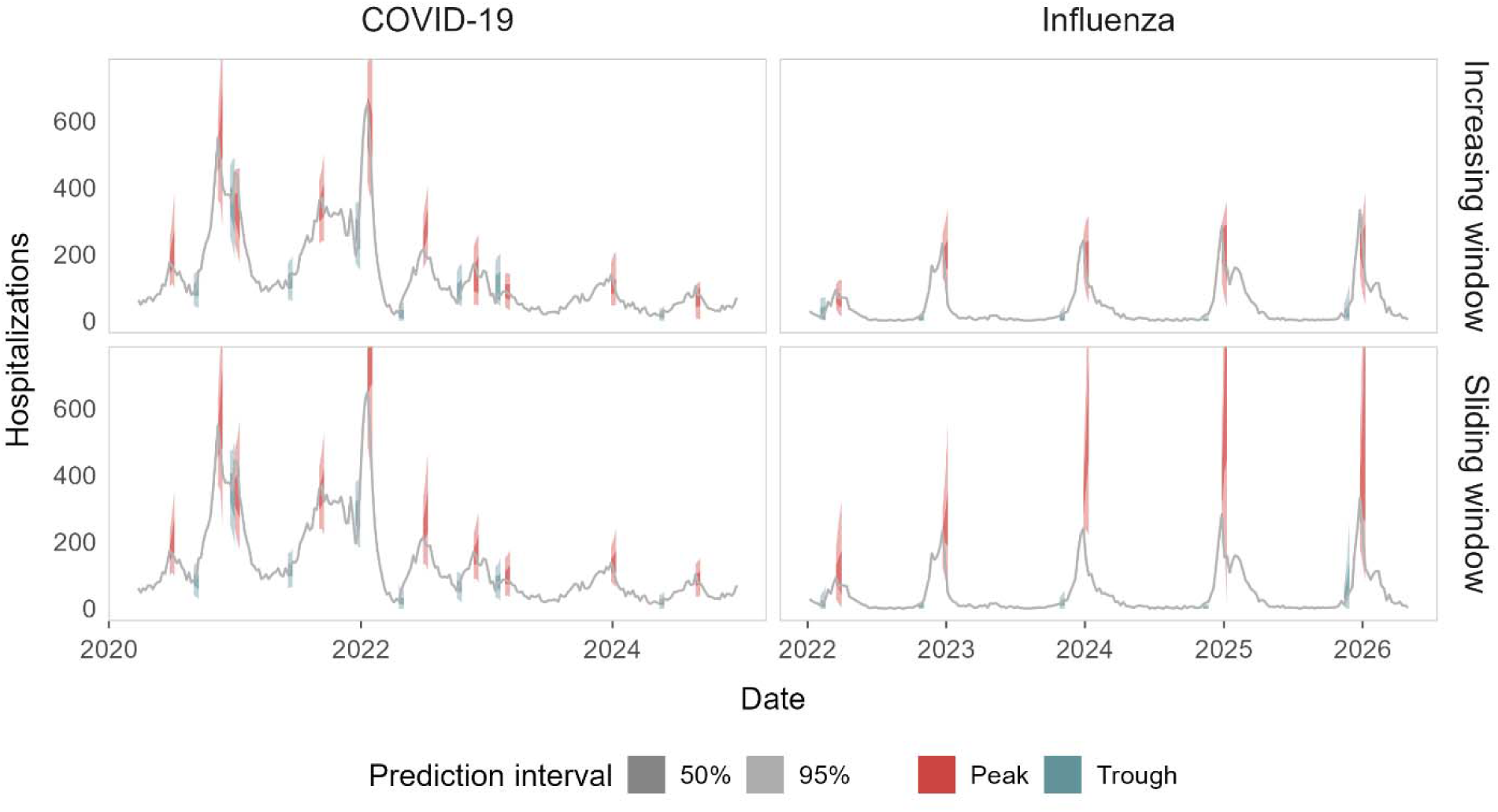
Peak and trough performance. Ensemble model (median of individual model types) 50% and 95% prediction intervals for 3-week hospitalization forecasts during peak and trough transmission periods. Influenza values prior to 2022 have been removed from the plot for visualization purposes. The dates of peak and trough periods are shown in **Figure S4**.

Most models did not consistently demonstrate strong performance across peaks and troughs in respiratory disease hospitalization trends (**Figure S12**). Across both pathogens, most models were ranked in both the top three best performers and the bottom three worst performers across peak and trough evaluation windows (COVID-19: 15/20, influenza: 15/20). Similarly, models under both training strategies were ranked in both the top three and bottom three across evaluation windows (increasing window: 14/20, sliding window: 17/20).

## DISCUSSION

Forecasting models often rely on the assumption that historical relationships between surveillance predictors and disease outcomes remain stable over time. However, respiratory disease systems are fundamentally nonstationary, with changing transmission dynamics, population immunity, circulating variants, and surveillance practices altering these relationships across seasons [14,15]. In this analysis of respiratory disease data streams across Utah, we found that the utility of syndromic surveillance and wastewater predictors was pathogen-, model-, and context-dependent. Forecast performance also varied substantially between increasing and sliding window training strategies and across periods of increasing, peak, and decreasing transmission. Together, these findings suggest that forecasting systems should prioritize flexibility, model diversity, and ongoing real-time evaluation of predictor utility.

Work in econometrics and environmental health has long recognized that pooling data across extended periods can bias cross-correlation functions and obscure lead-lag patterns occurring within narrower time frames [38]. This phenomenon was present in our Utah surveillance data, particularly for COVID-19, where relationships between hospitalizations and surveillance predictors varied considerably across dominant variant periods. When all data were analyzed together, adult and older adult predictors showed strong positive correlations with hospitalization counts, but this finding hid the weak or negative correlation patterns observed during the original SARS-CoV-2 strain and Alpha periods. Pooling data across long periods produces an aggregate cross-correlation function that may not accurately represent any individual period. Similar findings were observed nationally during the COVID-19 pandemic, when correlations and lead-lag relationships between wastewater and hospitalization, ED, and test positivity rates varied across variant surges in Houston [39] and pre- and post- U.S. Public Health Emergency periods in Maryland [40]. Shifts in wastewater correlations have been hypothesized to reflect changes in viral shedding dynamics over time due to variants and vaccination rates, while changes in ED and test positivity relationships may be driven by differences in disease severity, testing availability, healthcare-seeking behavior, and vaccination coverage [40].

In contrast to COVID-19, influenza surveillance streams showed stronger and more consistent positive correlations across age-stratified hospitalizations and predictors, likely reflecting broader transmission across age groups during seasonal influenza. However, pooling all years still obscured variation between seasons. Many influenza forecasting approaches implicitly assume that relationships between surveillance predictors and outcomes remain stable from year to year. Our findings suggest that this is not always the case, potentially due to differences in H1N1 and H3N2 dominance, although we lacked sufficient non-pandemic data to investigate this hypothesis. These findings emphasize the importance of continually evaluating surveillance predictors rather than relying on models and data streams that performed well in previous seasons.

In addition to temporal heterogeneity in predictor relationships, model performance also varied across time. GAMs with covariates that performed well during the Alpha and Delta SARS-CoV-2 waves performed worse than benchmark models in over half of forecasting windows during Omicron. Similarly, benchmark and time series models that performed well during peak influenza activity were often also among the worst performing models during troughs. Model rankings have likewise varied during annual CDC FluSight challenges, where top-performing models in one season do not consistently perform well in subsequent seasons [19,41]. The frequent reversal of model rankings suggests that ensemble approaches may derive their strength less from identifying the single best model and more from maintaining diversity [9] across model structures and covariates.

Diversity among model training strategies may also be important for ensembling. Forecasts generated under a sliding window strategy in our analysis generally outperformed those generated under an increasing window strategy, indicating that, contrary to the common assumption that including more historical data will improve forecasting performance, training models on more data actually resulted in reduced accuracy. The two exceptions were influenza models trained under an increasing window strategy which performed better during peaks, and Mantis models, which were robust to training methodology. These findings suggest that disease type and forecasting target (e.g., peak intensity prediction vs overall seasonal performance) should be considered when selecting a training strategy.

Limitations to our analysis include the inability to separate out the effects of tourism in wastewater predictors and to stratify hospitalizations and ED visits by influenza subtype. Because forecasts were generated retrospectively, we also did not account for lags in data-stream availability, including delays in data reporting from hospitals and sewersheds. However, surveillance data streams had high coverage, capturing 100% of hospitalizations and ED visits among Utah residents receiving care within the state, and an estimated 88% of the population through wastewater surveillance. A strong partnership between academia and government facilitated data sharing and creation of an adaptive forecasting workflow. Future directions include extending these methods to the sub-state level for Utah’s local health districts and evaluating the usefulness of state and local forecasts for decision-making during the 2026-2027 respiratory virus season.

Findings from this forecasting exercise highlight the challenges of forecasting respiratory disease hospitalizations in settings where surveillance predictors, healthcare-seeking behaviors, and pathogen characteristics evolve over time. Across both diseases, we found that the value of individual predictors, model classes, and training strategies varied substantially across seasons. These results indicate that forecasting pipelines should prioritize continual evaluation of predictor utility and diversify ensembles, rather than relying on any single model, surveillance stream, or historical training framework. Maintaining this flexibility also requires sustained partnerships between forecasters and public health agencies. Rather than building academic-government partnerships only during emergencies, programs that connect these two groups require sustained investment to maintain forecasting capacity between periods of heightened disease activity. While progress has been made in short-term respiratory disease forecasting, accurately forecasting hospitalization trends during times of rapid epidemiologic change, particularly hospitalization peaks, remains a major challenge. Future forecasting efforts will benefit from the ability to identify times when historical relationships between predictors and outcomes no longer hold so that models can be adapted accordingly.

## DATA AVAILABILITY

All code used in this study has been made publicly available on GitHub: https://github.com/EpiForeSITE/respiratory-forecasting-Utah. Shapefiles used in this study are publicly available from the Utah Geospatial Resource Center: https://gis.utah.gov. Aggregated respiratory disease surveillance data is publicly available from UDHHS: https://dhhs.utah.gov/health-dashboards/respiratory-disease-data/. Aggregated wastewater data is publicly available from UDHHS: https://avrpublic.dhhs.utah.gov/uwss/. Daily-level respiratory disease data was provided by UDHHS; access is restricted to protect patient confidentiality but can be made available upon request.

## AUTHOR CONTRIBUTIONS

HMT, TS, RG, and LTK contributed to project conceptualization. TS, RG, JK, and NL curated data. HT and TS conducted formal analysis and validation. HMT prepared the original draft manuscript and all authors contributed to review and editing.

## ACKNOWLEDGEMENTS

We are grateful for the support provided by Carson Dudley and Reiden Magdaleno for use of the Mantis model which they developed at the University of Michigan. We also thank Theron Jeppson and Emanuel Vasquez Gomez from the Division of Population Health Informatics Program at UDHHS for their help in setting up data pulls.

