## Supplemental Material for "Rethinking respiratory disease forecasting: temporal heterogeneity between surveillance predictors and outcomes drives forecast instability"

#### TITLE

### TECHNICAL METHODS

#### Models

We modeled forecasts using a suite of statistical and mechanistic-trained models:

*Naïve*: uses the most recently observed hospitalization count as the predicted forecast for all future time points, with uncertainty derived from the variance of historical errors.

*Drift*: an extension of the naïve model which incorporates a linear trend to forecasts based on the average change from the first to the most recent observation.

*Auto Regressive Integrated Moving Average (ARIMA)*: forecasts time-series data by using past hospitalization counts, trends, and errors to identify patterns and predict future values. Models combine autoregressive (lags) and moving average components (lagged errors) alongside a differencing step to remove trends.

*Error, Trend, Seasonal (ETS)*: decomposes time series data into values for error, trend, and seasonality with each component updating over time via exponential smoothing. Our models use additive errors and allow the model to choose the best values for trend and season (none, additive, multiplicative).

*Prophet*: a decomposable time series model developed by Meta with piecewise linear or logistic trends which can accommodate multiple seasonalities, change points, and holiday effects.[1]

*Generalized Additive Models (GAM)*: an extension of linear regression which uses spline-based non-linear smoothing functions to fit the effects of predictors and time. GAM models incorporated covariates from syndromic surveillance and wastewater data including all-age ED count, 65+ ED count, all-age percent of ED visits, 65+ percent of ED visits, and wastewater decile. GAM models used a negative binomial distribution to account for overdispersed count outcomes. These models included a smooth term for time and for one or more predictors. To improve model stability, the basis dimension for each smoothed covariate was capped at the number of unique observed minus one, with an upper boundary of 20. Forecast uncertainty for GAM models was generated using parametric Monte Carlo simulations. For each fitted model, we drew 2,000 samples of model coefficients from their estimated multivariate normal distribution. For each draw, we computed predicted hospitalization counts for the test data and simulated outcomes from a negative binomial distribution. Predictive intervals were obtained from the empirical quantiles of the simulated values.

*Mantis*: a foundation model developed at the University of Michigan trained entirely on mechanistic model simulations and generalized across disease types and outcomes with the ability to accommodate predictors.[2] Mantis models incorporated covariates from syndromic surveillance and wastewater data including all-age ED count, 65+ ED count, all-age percent of ED visits, and wastewater decile. An additional model was run with no covariates. The current version of Mantis (*rmantis v0.1.0*; Dudley and Magdaleno, 2026) accepts only a single covariate at a time and restricts inputs to cases, hospitalizations, and deaths. Mantis only accepts 112 weeks of training data per run, so the increasing window forecasting strategy limits data to this window. To accommodate these constraints, all predictors were input as cases and rescaled accordingly. Emergency department count and wastewater decile data were multiplied by 1,000, and emergency department percentages were multiplied by 10,000. Mantis operates on weekly inputs, and daily training data were aggregated to weekly values before model fitting.

#### Ensemble construction

Individual model forecasts were aggregated into an ensemble model for each disease and under each modeling method (increasing vs. sliding window). Ensembles were constructed using the median value for each time step's point forecast and for each prediction interval bound.

#### Converting daily forecasts to weekly forecasts

All models except Mantis were trained on daily data and generated daily forecasts. To convert daily forecasts to weekly-level forecasts, daily point predictions were summed. Weekly uncertainty intervals were derived using a parametric bootstrap simulation approach that approximated daily standard errors from prediction intervals and combined daily variances while accounting for correlation between days using a fixed correlation parameter. Weekly 50% and 95% intervals were estimated from the empirical quantiles of simulated weekly samples.

**FIGURES AND TABLES**

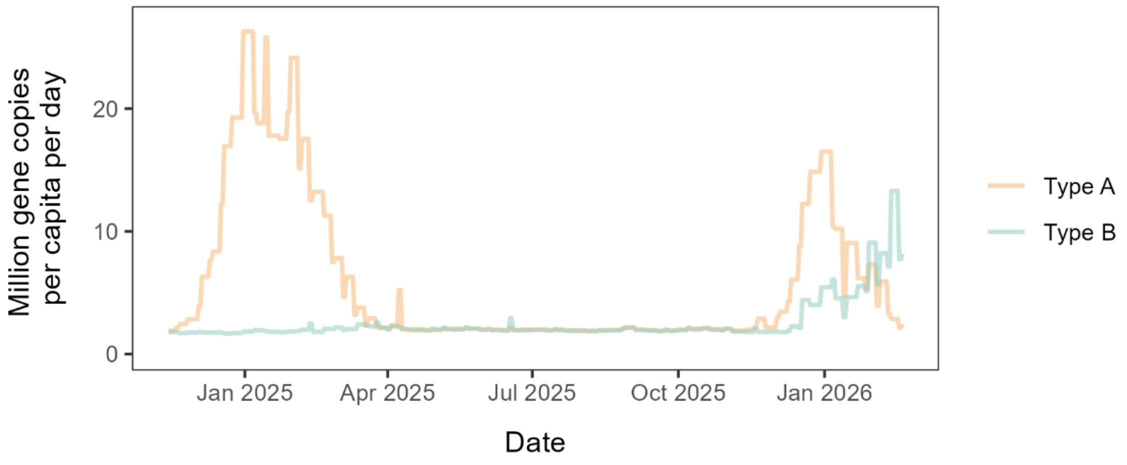


**Figure S1**. **Wastewater influenza sub-types**. Influenza viral load (million gene copies per capita per day) collected from wastewater. Samples are generally collected twice per week from Utah's 35 wastewater collection sites. Values are reported daily and held constant from the last sample collection until the next sample is collected. Wastewater detection levels of Influenza subtype A were used in the primary analysis.

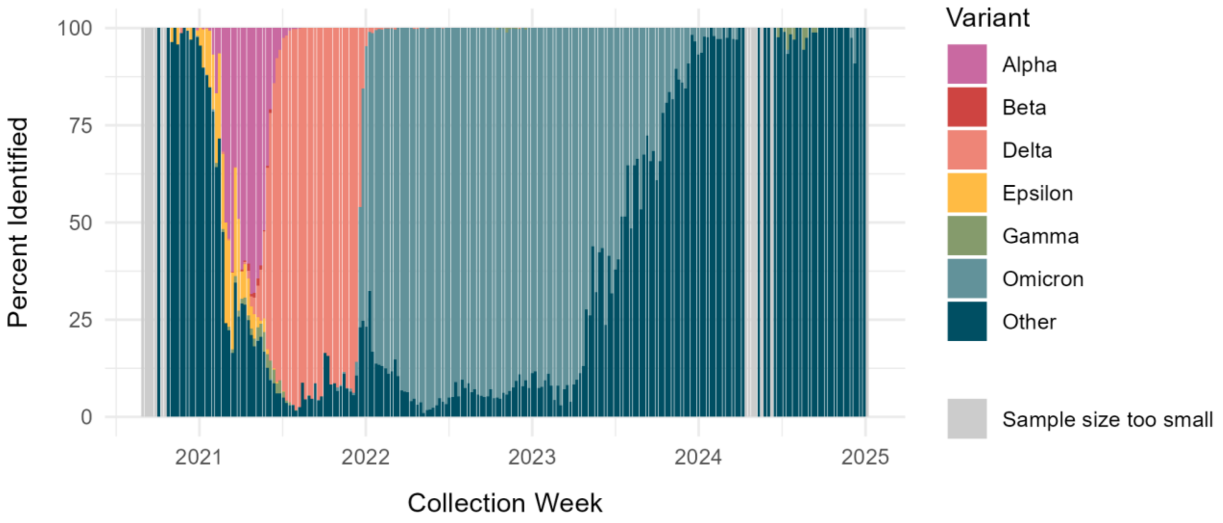


**Figure S2**. **SARS-CoV-2 variant distribution**. SARS-CoV-2 variant sequencing results by week. Sequences are derived from clinical samples. Results from weeks with less than 20 sequences reported are censored.

**Table S1**. **Models**. Model types and associated predictors used to forecast respiratory diseases in Utah.

| *Model* | *Covariates* | | | | | |
|---|---|---|---|---|---|---|
| | *None* | *All-age ED count* | *65+ ED count* | *All-age ED %* | *65+ ED %* | *Wastewater decile* |
| *Naïve* | x | | | | | |
| *Drift* | x | | | | | |
| *ARIMA* | x | | | | | |
| *ETS* | x | | | | | |
| *Prophet* | x | | | | | |
| *GAM1* | | x | | | | |
| *GAM2* | | | x | | | |
| *GAM3* | | | | x | | |
| *GAM4* | | | | | x | |
| *GAM5* | | | | | | x |
| *GAM6* | | x | | | x | |
| *GAM7* | | | x | x | | |
| *GAM8* | | x | | | x | x |
| *GAM9* | | | x | x | | x |
| *Mantis1* | x | | | | | |
| *Mantis2* | | x | | | | |
| *Mantis3* | | | x | | | |
| *Mantis4* | | | | x | | |
| *Mantis5* | | | | | x | |
| *Mantis6* | | | | | | x |

**Note**: ED = emergency department, ARIMA = Auto Regressive Integrated Moving Average, ETS = Error, Trend, Seasonality

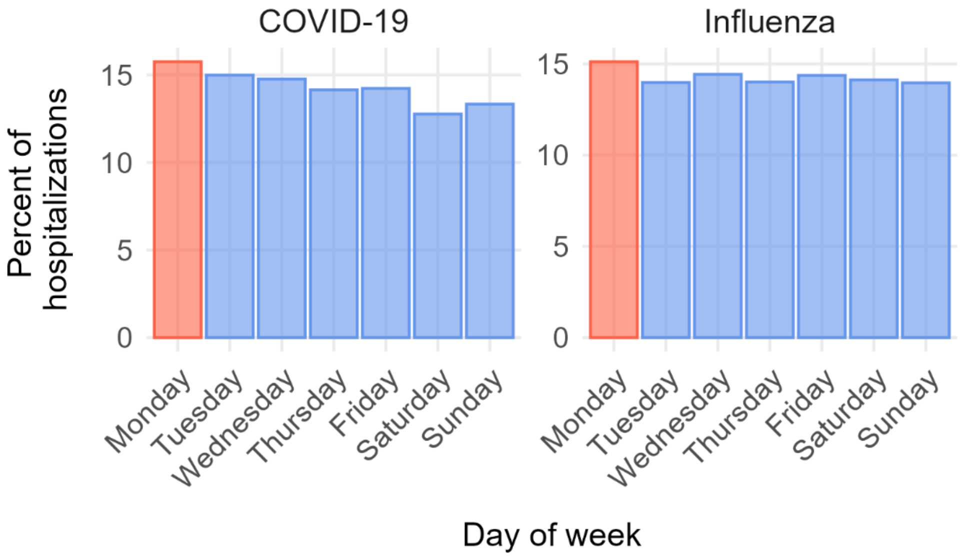


**Figure S3**. **Weekly hospitalization breakdown**. Percent of respiratory disease hospitalizations occurring on each day of the week across the state of Utah. COVID-19 data are from March 20, 2020, to May 1, 2026, and influenza data are from January 1, 2018, to May 1, 2026.

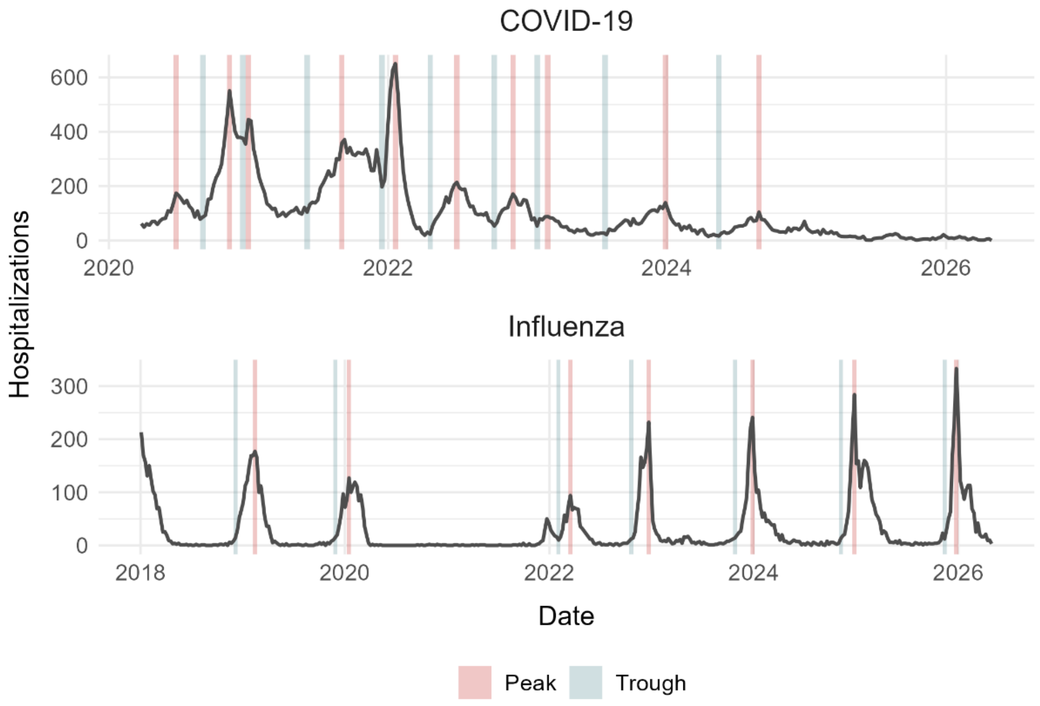


**Figure S4**. **Peaks and troughs**. Peak (high transmission) and trough (low or no transmission) periods of rapid change identified in respiratory disease hospitalization trends.

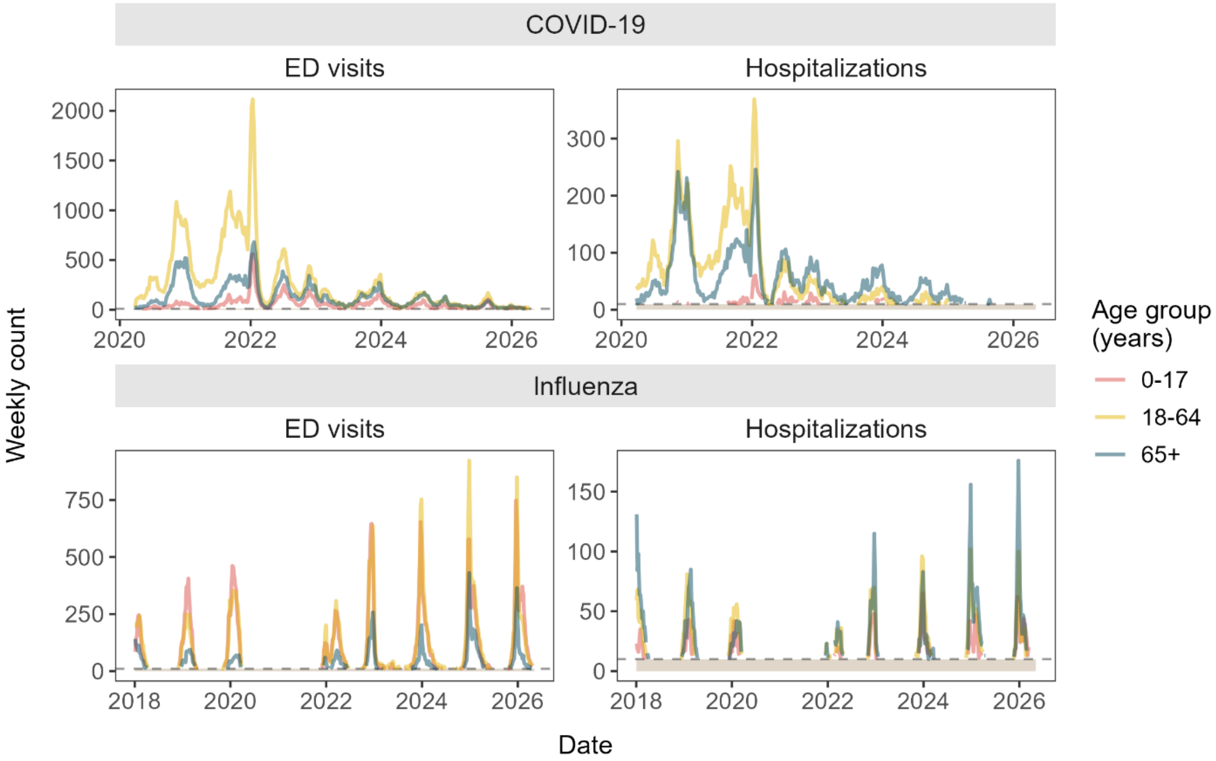


**Figure S5**. **Age-stratified data streams**. Hospitalizations and emergency department (ED) visits, stratified by infection and age group. Counts between 1 and 10 per week are censored to protect patient confidentiality.

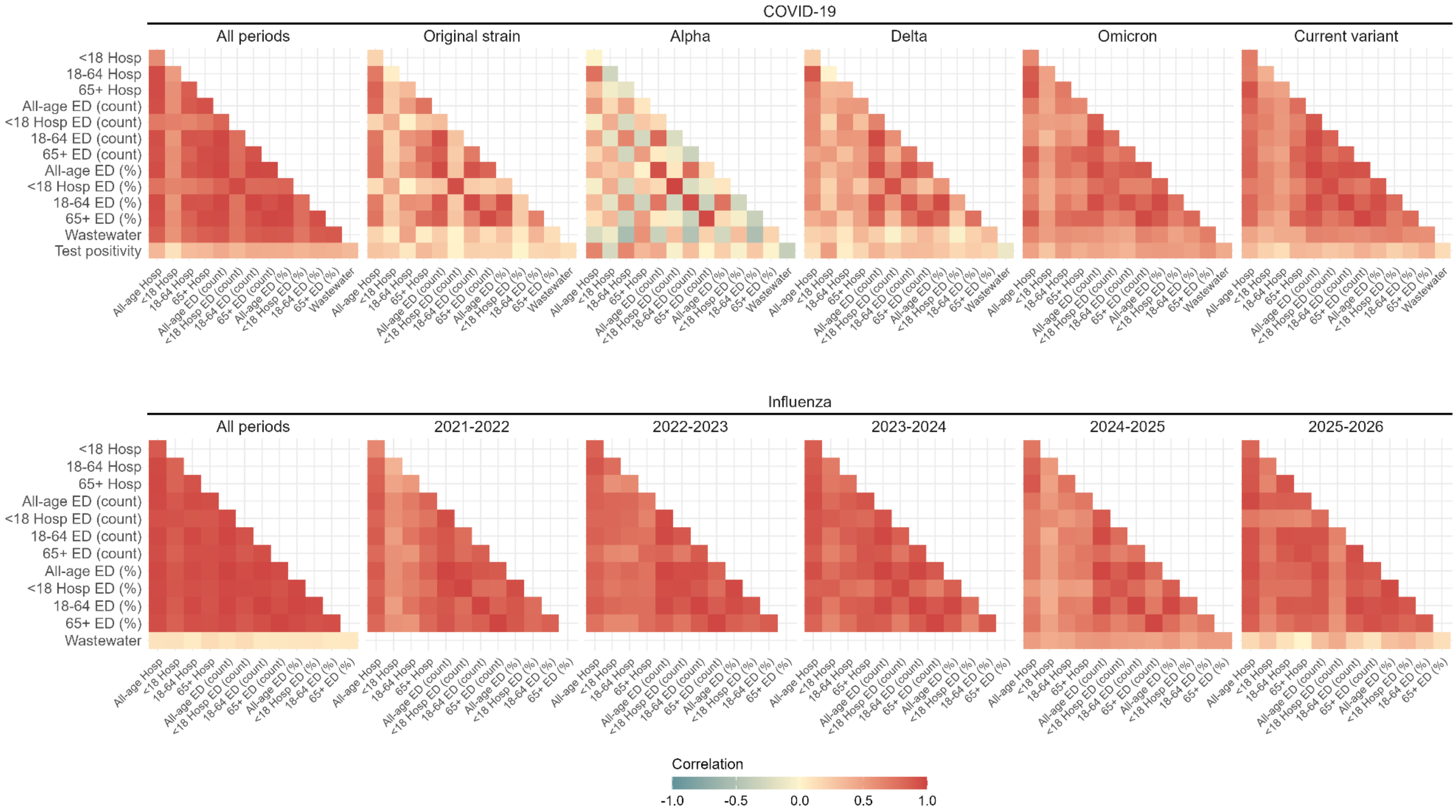


**Figure S6**. **Predictor cross-correlations**. Cross-correlation matrices of age-stratified hospitalization, emergency department (ED), and wastewater surveillance predictors for COVID-19 and influenza across all periods and by season or dominant variant. Warmer colors indicate stronger positive correlations, while cooler colors indicate weaker or negative correlations.

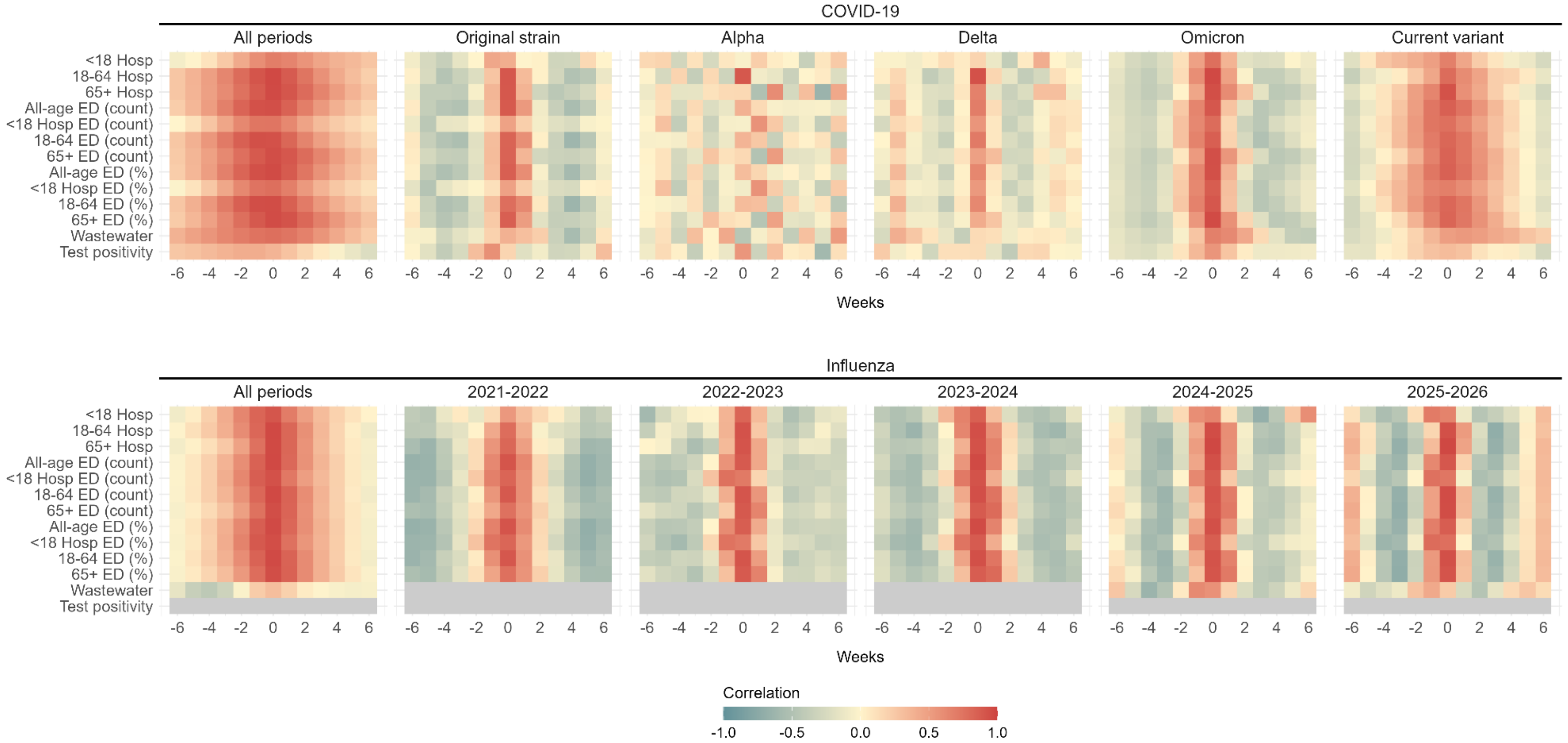


**Figure S7**. **Lagged cross-correlations**. Lagged correlations between all-age hospitalization counts and hospitalization, emergency department (ED) surveillance, wastewater, and test positivity (SARS-CoV-2 only) predictors. Results are stratified by COVID-19 and influenza across all periods and by season or dominant variant. Correlations are shown across weekly lags (-6 to +6 weeks), where negative lags indicate that predictors lag hospitalizations and positive lags indicate that predictors lead hospitalizations. Warmer colors indicate stronger positive correlations, while cooler colors indicate weaker or negative correlations.

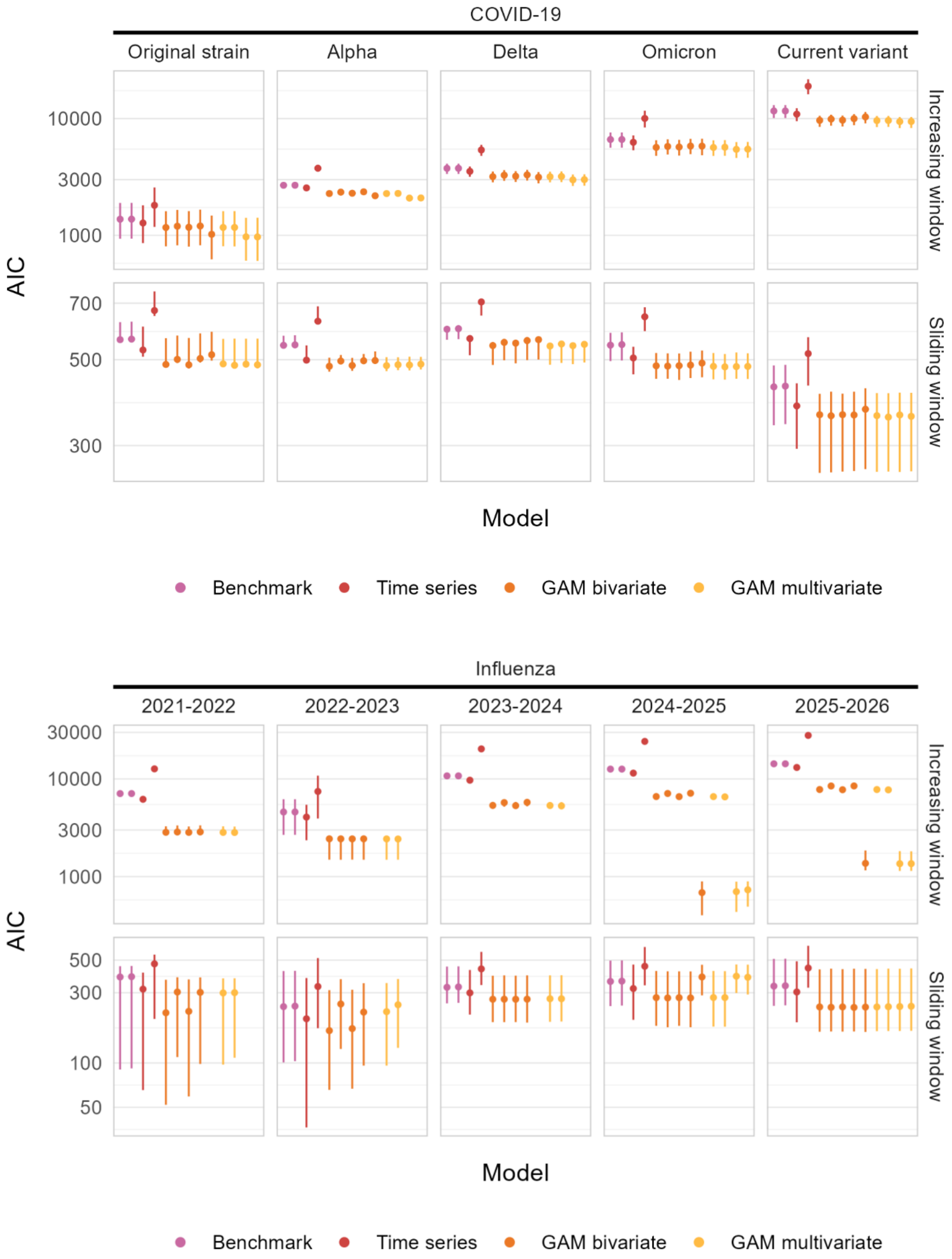


**Figure S8**. **AIC**. Median (interquartile range) Akaike Information Criterion (AIC) metrics for respiratory disease forecasts, stratified by season and time-series forecasting method. Note: GAM = generalized additive model

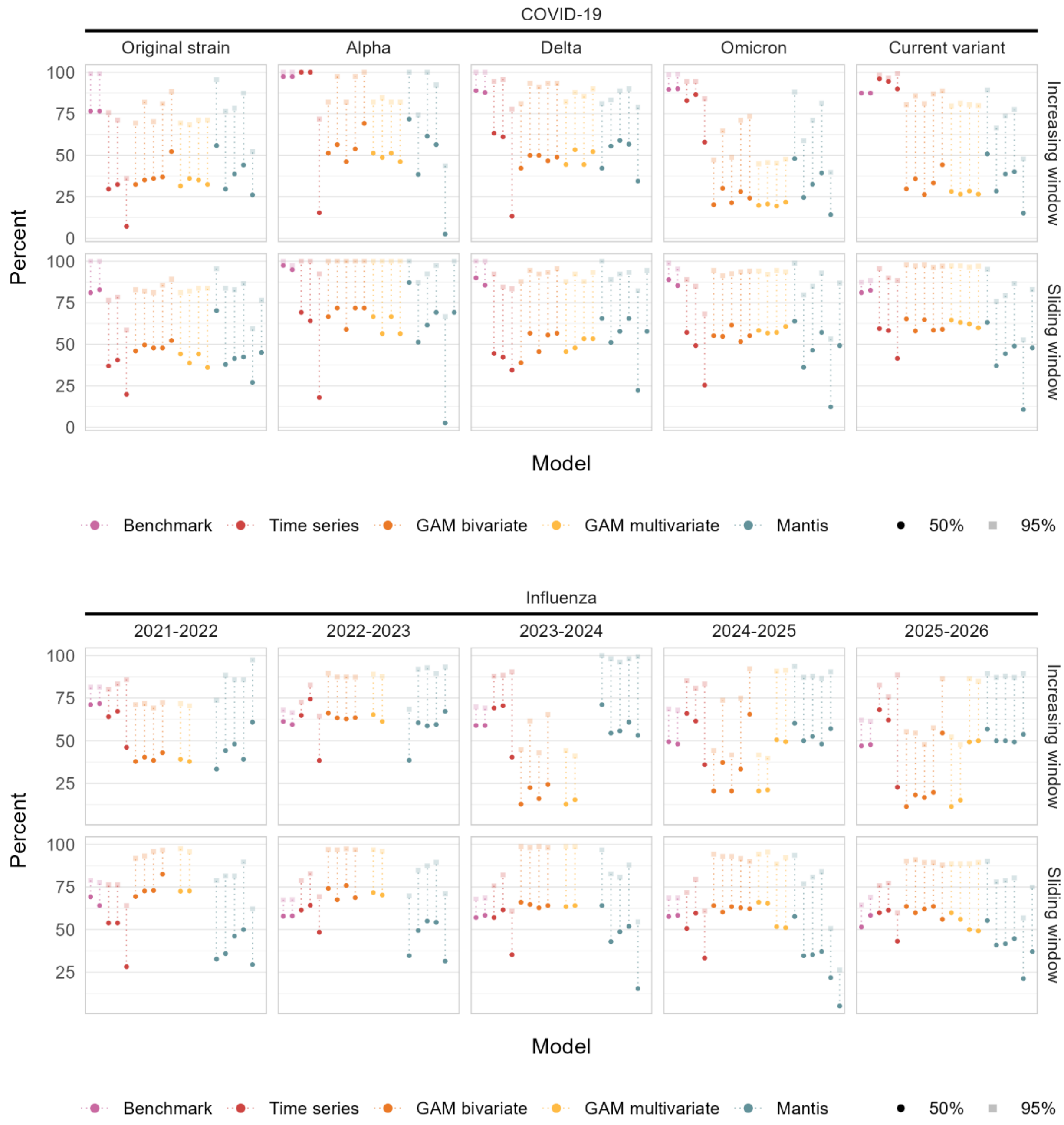


**Figure S9**. **Prediction interval coverage.** Prediction interval metrics (50% and 95%) for respiratory disease forecasts, stratified by season and time-series forecasting method. Note: GAM = generalized additive model

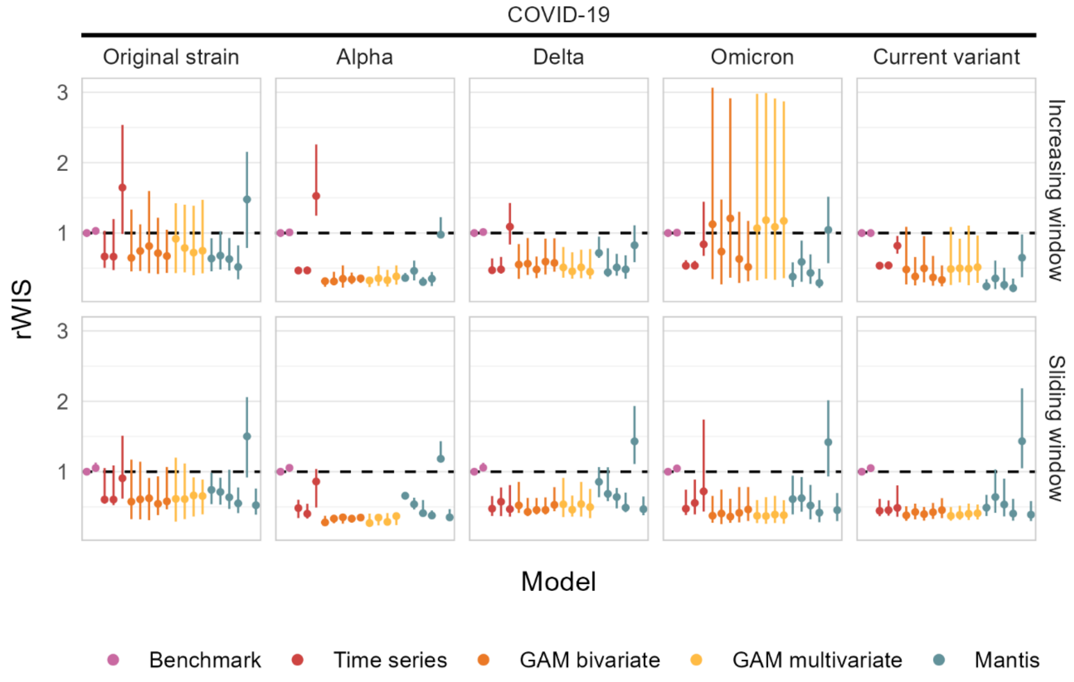


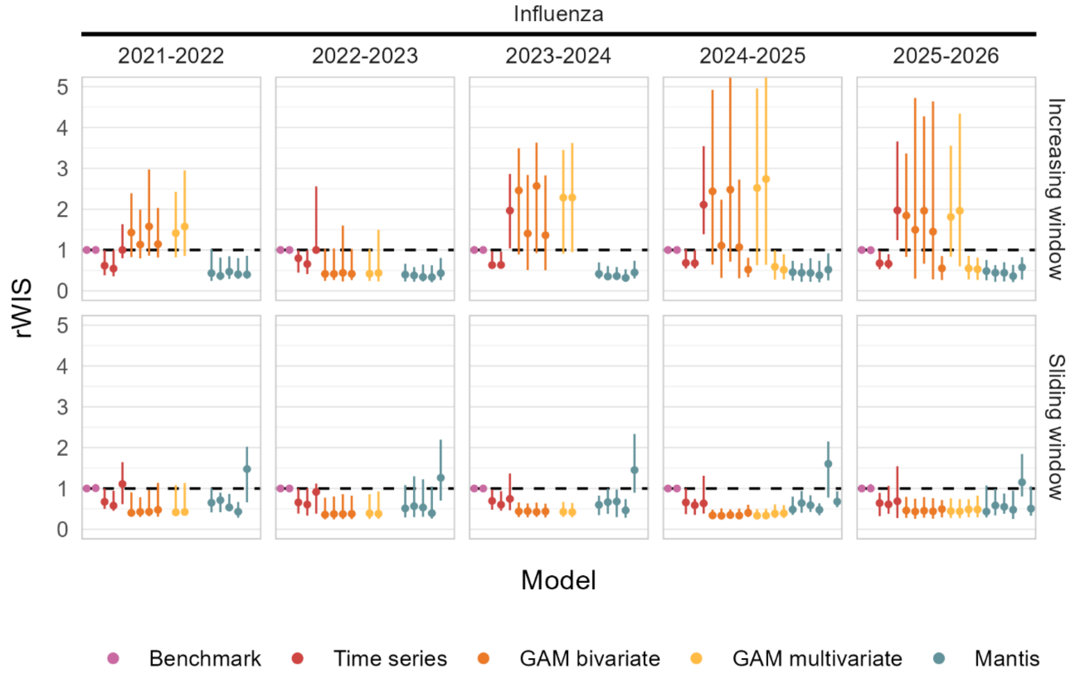


**Figure S10**. **Relative weighted interval score.** Median (interquartile range) relative weighted interval score (rWIS) metrics, stratified by season and time-series forecasting method. The dashed horizonal line indicates an rWIS value of 1, relative to a Naïve benchmark. Infinite values due to benchmark model WIS = 0 have been removed. Note: GAM = generalized additive model

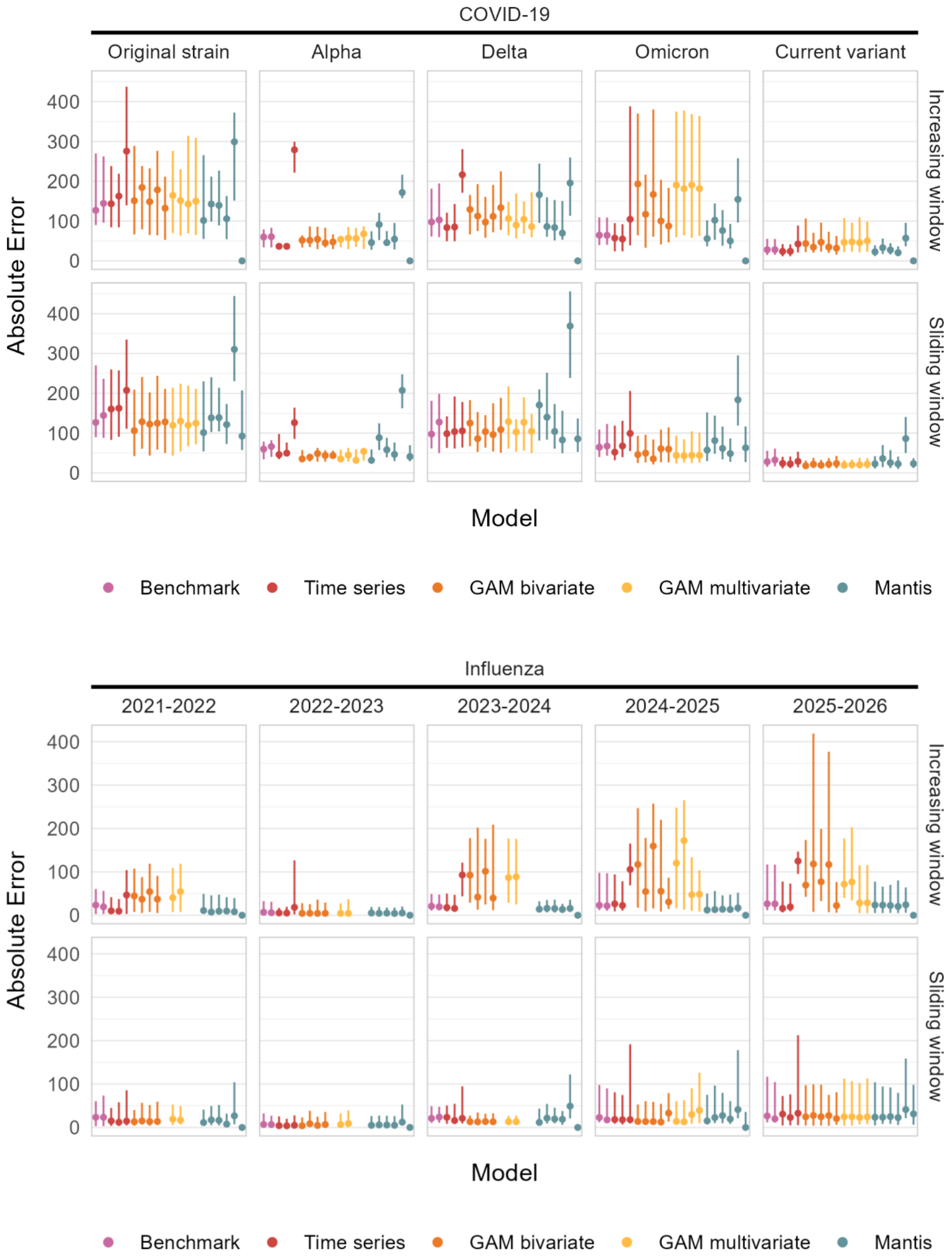


**Figure S11**. **Absolute error**. Median (interquartile range) absolute error metrics for respiratory disease forecasts, stratified by season and time-series forecasting method. Note: GAM = generalized additive model

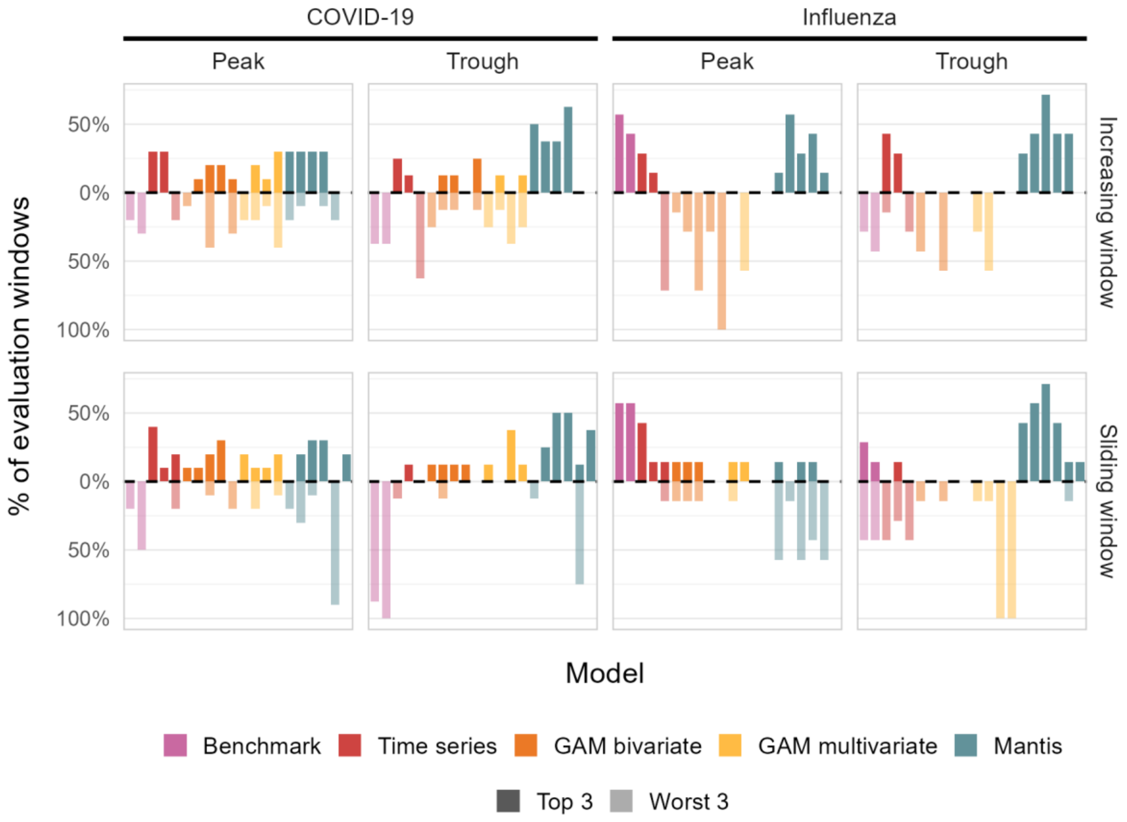


**Figure S12**. **Model rankings**. Frequency of top three and bottom three ranked model performance during respiratory hospitalizations peak and trough periods. Bars show the percentage of peak or trough evaluation windows in which each model ranked among the three best-performing models or three worst-performing models based on relative weighted interval score (rWIS). Positive values indicate the percentage of windows in which a model was ranked in the top three, and negative values indicate the percentage of windows in which a model was ranked in the bottom three. Results are stratified by disease and forecast training strategy. Bar colors indicate model class. The dashed horizontal line marks 0%, where models were neither frequently among the best nor worst performers.
